# COVID-19 Antibody in Thai Community Hospitals

**DOI:** 10.1101/2020.06.24.20139188

**Authors:** Tanawin Nopsopon, Krit Pongpirul, Korn Chotirosniramit, Narin Hiransuthikul

## Abstract

**Background:** COVID-19 seroprevalence data has been scarce, especially in less developed countries with a relatively low infection rate.

**Methods:** A locally developed rapid IgM/IgG test kit was used for screening hospital staff and patients who required procedural treatment or surgery in 52 hospitals in Thailand from April 8 to June 26, 2020. A total of 857 participants were tested—675 were hospital staff and 182 were pre-procedural patients. (Thai Clinical Trials Registry: TCTR20200426002)

**Results:** Overall, 5.5% of the participants (47 of 857) had positive immunoglobulin M (IgM), 0.2% (2 of 857) had positive immunoglobulin G (IgG) and IgM. Hospitals located in the Central part of Thailand had the highest IgM seroprevalence (11.9%). Preprocedural patients had a higher rate of positive IgM than the hospital staff (12.1% vs. 3.7%). Participants with present upper respiratory tract symptoms had a higher rate of positive IgM than those without (9.6% vs. 4.5%). Three quarters (80.5%, 690 of 857) of the participants were asymptomatic, of which, 31 had positive IgM (4.5%) which consisted of 20 of 566 healthcare workers (3.5%) and 11 of 124 preprocedural patients (8.9%).

**Conclusions:** COVID-19 antibody test could detect a substantial number of potential silent spreaders in Thai community hospitals. Antibody testing should be encouraged for mass screening, especially in asymptomatic individuals.

## Introduction

Polymerase chain reaction (PCR) was introduced as a diagnostic test of choice for coronavirus disease 2019 (COVID-19) infection. However, it might not be readily available or affordable in many facilities and could pose an unnecessary risk to the healthcare providers during the specimen collection. Besides, a recent study raised a concern of false-negative results from the PCR test for severe acute respiratory syndrome coronavirus 2 (SARS-CoV-2) in patients with high pretest probability and encouraged the development of a highly sensitive test [1]. In Thailand, the PCR test was offered in suspected individuals with strict criteria during the initial phase of the COVID-19 pandemic. As a more feasible, cheaper, and safer alternative to the PCR, the antibody test is not only useful for an epidemiological investigation [2] but could also be used for mass screening of potential silent spreaders— asymptomatic COVID-19 individuals.

Hospital is one of the best venues for getting and spreading pathogens. There are two types of people in the hospital who potentially are silent spreaders and need antibody testing: (1) healthcare workers who have a relatively higher risk of infection than laypersons, and (2) asymptomatic patients who need procedural treatment or operation but do not meet the criteria for PCR testing.

## Methods

From April 8 to June 26, 2020, hospital staff and patients who needed procedural treatment or operation but did not meet the national PCR testing criteria in 244 hospitals (215 community hospitals and 29 general hospitals) from all regions of Thailand were offered antibody testing. Of 215 community hospitals, data from 52 hospitals (24.2%) in 35 provinces were readily available for the analysis performed on June 29, 2020. Patients with PCR confirmed COVID-19 infection were quarantined and excluded. Baiya Rapid COVID-19 IgG/IgM test kit (Baiya Phytopharm, Thailand) which reports the presence of immunoglobulin M (IgM) and immunoglobulin G (IgG) qualitatively, was used in this study free of charge. The internal validation of the test kit using the serum of 51 PCR confirmed COVID-19 cases and 150 controls showed sensitivity 94.1% (48 of 51) and specificity 98.0% (147 of 150) for IgM or IgG antibody. Participants with positive IgM were encouraged to have a PCR test if available.

### Age-adjusted seroprevalence

PCR confirmed COVID-19 cases were obtained from the Thailand government report on June 30, 2020. Seroprevalence data were presented as unadjusted seroprevalence and compared with direct age-adjusted seroprevalence using combined participating population, Thailand population, and world population provided by world health organization (WHO) for 2000-2025 population.

### Statistical analysis

Categorical data were presented with counts and percentages while continuous data were provided with median and interquartile range. The 95% confidence interval (CI) of the seroprevalence was calculated by Wilson’s method using binomial probabilities. Correlation between seroprevalence and PCR confirmed COVID-19 prevalence was tested using Spearman’s correlation. Missing data were excluded. A two-tailed p < 0.05 was considered statistically significant. All data were analyzed using Stata 16.1 (College Station, TX).

### Ethics Committee Approval

This study was approved by the Institutional Review Board of Chulalongkorn University (IRB No.236/63) and 18 general hospitals. Given no ethics committees were available in the participating community hospitals, participation in this study was approved by the hospital directors or representatives. All participants provided written informed consent. (Thai Clinical Trials Registry: TCTR20200426002)

## Results

Overall, 52 community hospitals from 46.1% of provinces in Thailand (35 of 76) which consisted of 58.2% of national population (35,416,545 of 60,892,671) participated in this study. Participation rates varied across regions— Northeastern (55%), Central (50.0%), Southern (42.9%), Northern (33.3%), and Eastern (28.6%) [Table 1].

**Table 1.** Geographical Distribution of 52 Participating Community Hospitals

|  | Participating<br>Provinces | Total<br>Provinces | Population of<br>Participating Provinces | Population of<br>All Provinces |
| --- | --- | --- | --- | --- |
| Thailand* | 35 (46.1%) | 76 | 35,416,545 (58.2%) | 60,892,671 |
| Northern | 3 (33.3%) | 9 | 2,505,118 (39.4%) | 6,350,499 |
| Northeastern | 11 (55.0%) | 20 | 15,064,919 (68.4%) | 22,014,248 |
| Central* | 13 (50.0%) | 26 | 10,053,397 (55.3%) | 18,192,361 |
| Southern | 6 (42.9%) | 14 | 5,697,112 (60.0%) | 9,493,757 |
| Eastern | 2 (28.6%) | 7 | 2,095,999 (43.3%) | 4,841,806 |

\* Not include Bangkok which has no community hospital.
Data were presented in counts and percentages.

From 52 community hospitals, 857 participants which consisted of 675 hospital staff and 182 pre-procedural patients were included in the study. Their median age was 37 years (interquartile range 27–45), 74.7% were female, 98.8% were Thai, and 80.5% were asymptomatic. The most common symptoms were cough (9.7%), rhinitis (7.5%), sore throat (6.4%), fever (5.7%), and dyspnea (3.5%). History of travel to the high-risk area was 6.0%, history of close contact to the confirmed COVID-19 case was 15.4%, and 14.5% had PCR negative. Forty-seven participants (5.5%, 95% CI 4.1–7.2) had IgM antibody against SARS-CoV-2 whereas the IgG antibody was found in two participants (0.2%, 95% CI 0.1–0.8). Participants from the Central region of Thailand had the highest IgM seroprevalence (11.9%, 95% CI 8.4–16.5), while the Northern region had the lowest seroprevalence (1.6%, 95% CI 0.3–8.7) [Fig 1].

**Fig 1.**
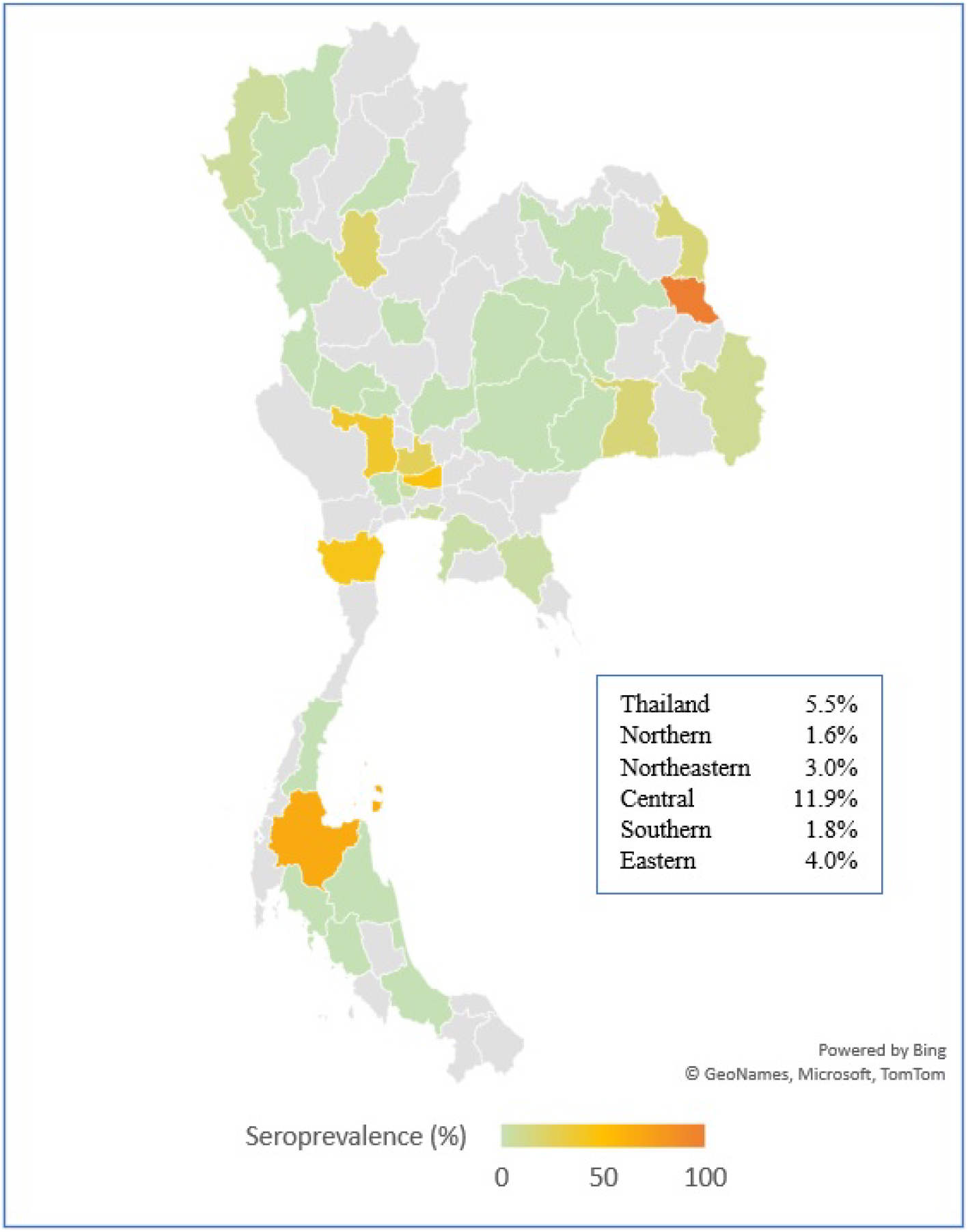
Unadjusted IgM Seroprevalence in Community Hospitals across Geographical Regions of Thailand.

Age-adjusted IgM seroprevalence with combined participating population showed almost similar results with unadjusted IgM seroprevalence. However, age-adjusted seroprevalence using Thailand population showed increasing seropositive rate in Thailand from 5.5% to 6.3%, Central region from 11.9% to 15.3%, and Northern region from 1.6% to 1.8%, while decreasing seroprevalence in Northeastern region from 3.0% to 1.6%, Eastern region from 4.0% to 2.8%, and Southern region from 1.8% to 1.5%. Adjusted with the world standard population from the World Health Organization (WHO 2000-2025) showed decreasing trends of seroprevalence both overall and in most regions [Table 2].

**Table 2.** Unadjusted and age-adjusted immunoglobulin M seroprevalence in community hospitals across geographical regions of Thailand

|  | Unadjusted IgM<br>Seroprevalence | Age-adjusted IgM<br>Seroprevalence with Combined<br>Participating Population | Age-adjusted IgM<br>Seroprevalence with<br>Thailand Population | Age-adjusted IgM<br>Seroprevalence with World<br>Standard Population |
| --- | --- | --- | --- | --- |
| Thailand* | 5.5% | NA | 6.3% | 5.1% |
| Northern | 1.6% | 2.2% | 1.8% | 2.1% |
| Northeastern | 3.0% | 3.0% | 1.6% | 1.5% |
| Central* | 11.9% | 11.9% | 15.3% | 12.2% |
| Southern | 1.8% | 2.4% | 1.5% | 1.2% |
| Eastern | 4.0% | 3.9% | 2.8% | 2.6% |

\* Not include Bangkok which has no community hospital. IgM, immunoglobulin M; NA, not available; WHO, World Health Organization.

Pre-procedural patients had an unexpectedly higher proportion of positive IgM than the hospital staff (12.1% vs. 3.7%), especially patients in the Central region of Thailand (27.9%, 95% CI 18.2–40.2) while patients in the Northern and Southern regions showed zero seroprevalences. Also, hospital staff in the Central region had the highest seroprevalence (6.6%, 95% CI 3.8–11.1) while those in the Northern region had the lowest (1.7%, 95% CI 0.3–9.0). Overall, the seropositive prevalence was not different between males and females (5.8% vs. 5.5%).

Paradoxically, the seroprevalences were higher in participants without a history of travel to the high-risk area (5.6% vs. 3.9%) and those without a history of close contact to confirmed COVID-19 case (5.7% vs. 4.5%) than their counterparts. The same paradox also applied to pre-procedural patients. Patients without travel history were likely to have an antibody for SARS-CoV-2 (13.9% vs. 3.2%) and patients without close contact to the case also had more chance to develop an antibody (12.3% vs. 9.1%). However, healthcare workers with travel history had slightly more chance to develop IgM (5.0% vs. 3.7%) and with close contact history (4.1% vs. 3.6%). In general, participants with upper respiratory tract symptoms had a higher chance of being seropositive (9.6% vs. 4.5%), of which dyspnea had the highest (30.0%, 95% CI 16.7–47.9). Likewise, pre-procedural patient with dyspnea had the most IgM positive (29.6%, 95% CI 15.9–48.5) and healthcare worker with dyspnea (33.3%, 95% CI 6.1–79.2). Of 690 participants without present upper respiratory tract symptom, 31 had IgM positive for COVID-19 (4.5%, 95% CI 3.2–6.3) which consisted of 20 of 566 healthcare workers (3.5%, 95% CI 2.3–5.4) and 11 of 124 patients (8.9%, 95% CI 5.0–15.2). History of negative PCR was associated with a surprisingly higher chance of seropositive than those with no PCR test result (6.5% vs. 5.3%) [Table 3]. Unfortunately, none of participants with positive IgM had opportunities for PCR testing.

**Table 3.** Demographic Characteristics and Seroprevalence in different groups

|  | All Participants |  | Hospital Staff |  | Pre-procedural Patients |  |
| --- | --- | --- | --- | --- | --- | --- |
|  | n | IgM+ | n | IgM+ | n | IgM+ |
| Total | 857 | 47 (5.5%) | 675 | 25 (3.7%) | 182 | 22 (12.1%) |
| Median age, years<br>(25th–75th percentile) | 37 (27–45) |  | 36.5 (28–45) |  | 37 (25–53) |  |
| Male, n (%) | 207 (24.1%) | 12 (5.8%) | 145 (21.5%) | 6 (4.1%) | 62 (34.1%) | 6 (9.7%) |
| Female, n (%) | 640 (74.7%) | 35 (5.5%) | 521 (77.2%) | 19 (3.6%) | 119 (65.4%) | 16 (13.4%) |
| Unspecified, n (%) | 10 (1.2%) | 0 (0.0%) | 9 (1.3%) | 0 (0.0%) | 1 (0.5%) | 0 (0.0%) |
| Thai | 847 | 47 (5.5%) | 671 | 25 (3.7%) | 176 | 22 (12.5%) |
| Non-Thai | 10 | 0 (0.0%) | 4 | 0 (0.0%) | 6 | 0 (0.0%) |
| Region |  |  |  |  |  |  |
| North | 61 | 1 (1.6%) | 59 | 1 (1.7%) | 2 | 0 (0.0%) |
| Northeast | 269 | 8 (3.0%) | 220 | 4 (1.8%) | 49 | 4 (8.2%) |
| Central | 244 | 29 (11.9%) | 183 | 12 (6.6%) | 61 | 17 (27.9%) |
| South | 109 | 2 (1.8%) | 88 | 2 (2.3%) | 21 | 0 (0.0%) |
| East | 174 | 7 (4.0%) | 125 | 6 (4.8%) | 49 | 1 (2.0%) |
| History of travel to high risk area |  |  |  |  |  |  |
| Yes | 51 | 2 (3.9%) | 20 | 1 (5.0%) | 31 | 1 (3.2%) |
| No | 806 | 45 (5.6%) | 655 | 24 (3.7%) | 151 | 21 (13.9%) |
| History of close contact confirmed case |  |  |  |  |  |  |
| Yes | 132 | 6 (4.5%) | 121 | 5 (4.1%) | 11 | 1 (9.1%) |
| No | 725 | 41 (5.7%) | 554 | 20 (3.6%) | 171 | 21 (12.3%) |
| Asymptomatic | 690 | 31 (4.5%) | 566 | 20 (3.5%) | 124 | 11 (8.9%) |
| Symptomatic | 167 | 16 (9.6%) | 109 | 5 (4.6%) | 58 | 11 (19.0%) |
| Fever | 49 | 7 (14.3%) | 17 | 0 (0.0%) | 32 | 7 (21.9%) |
| Cough | 83 | 8 (9.6%) | 52 | 1 (1.9%) | 31 | 7 (22.6%) |
| Rhinitis | 64 | 7 (10.9%) | 47 | 2 (4.3%) | 17 | 5 (29.4%) |
| Sore throat | 55 | 8 (14.5%) | 37 | 3 (8.1%) | 18 | 5 (27.8%) |
| Dyspnea | 30 | 9 (30.0%) | 3 | 1 (33.3%) | 27 | 8 (29.6%) |
| Previous PCR status |  |  |  |  |  |  |
| Negative | 124 | 8 (6.5%) | 77 | 1 (1.3%) | 47 | 7 (14.9%) |
| Never tested | 733 | 39 (5.3%) | 598 | 24 (4.0%) | 135 | 15 (11.1%) |

Data were presented in counts and percentages unless otherwise specified.
IgM+, immunoglobulin M positive; NA, not available; PCR, polymerase chain reaction.

PCR confirmed COVID-19 case data were acquired for participating provinces with population. Overall, COVID-19 prevalence was 2.44 cases per 100,000. Participating provinces in the Eastern region had the highest prevalence of COVID-19 (6.54 cases per 100,000) while provinces in northeastern had the lowest prevalence [Table 4]. There was no correlation between IgM seroprevalence and PCR confirmed COVID-19 prevalence (p=0.199).

**Table 4.**
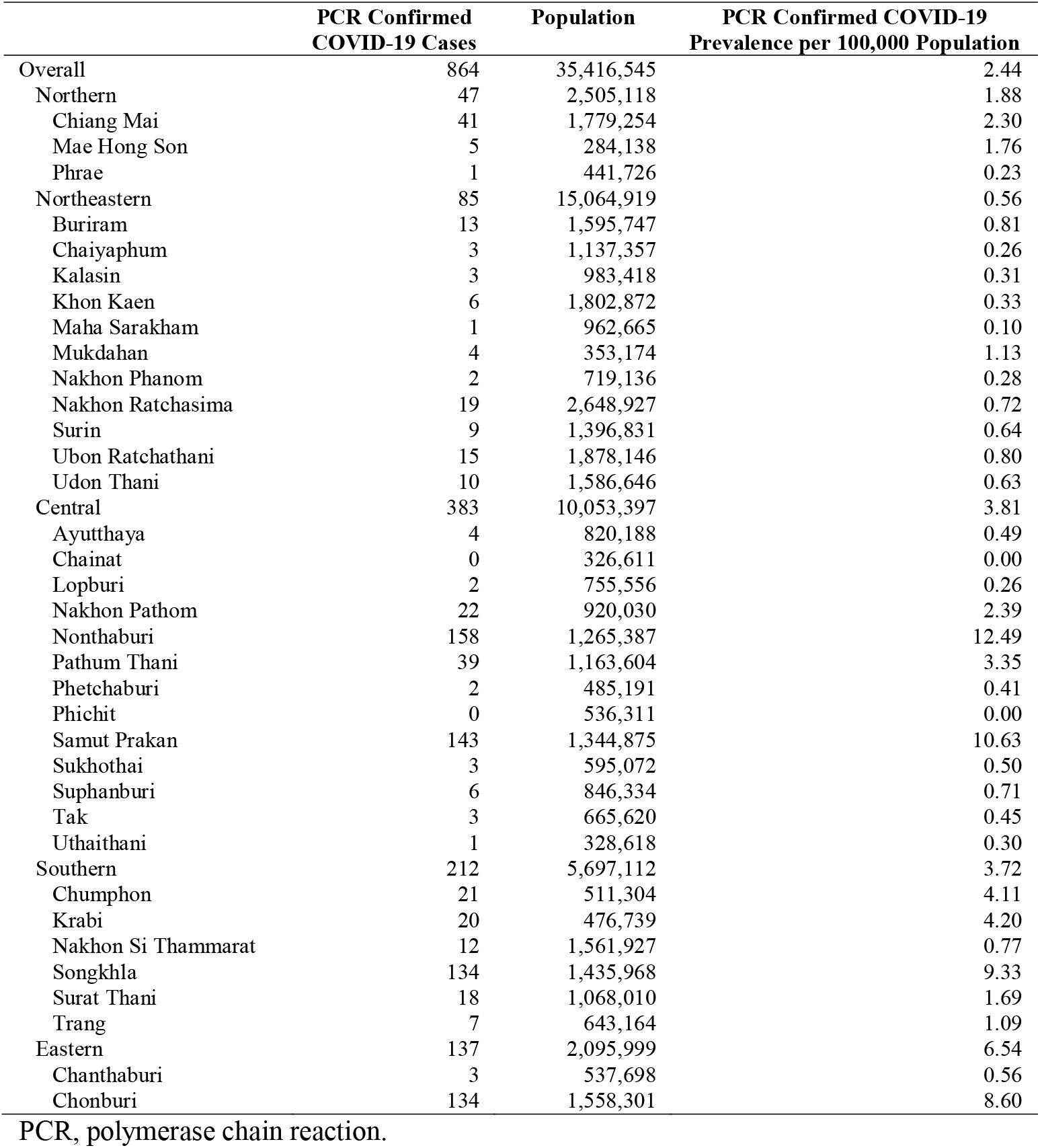
Number and prevalence of PCR confirmed COVID-19 cases in participating provinces

IgG was detected in two participants (0.2%, 95% CI 0.1–0.8) who also had a positive IgM antibody. In other words, we did not find any participants with isolated positive IgG. Participant A was a Thai female healthcare worker who worked in a community hospital in the Central region of Thailand. She had a sore throat but had no history of travel to a high-risk area or close contact to confirmed COVID-19 case, and did not have a PCR test before. Participant B was a Thai female preprocedural patient who visited another community hospital in the Central region. She had no symptom, no history of travel to a high-risk area, or close contact to confirmed COVID-19 case. Patient B had a previously negative PCR result [Table 5].

**Table 5.** Characteristics of Participants who Developed Immunoglobulin G Antibody

|  | Age Range, years | Gender | Ethnicity | Region | Occupation | History of travel to high-risk area | History of contact confirmed case | Symptoms | Previous PCR status |
| --- | --- | --- | --- | --- | --- | --- | --- | --- | --- |
| Participant A | 56-60 | Female | Thai | Central | HCW | No | No | Sore throat | Never tested |
| Participant B | 41-45 | Female | Thai | Central | Patient | No | No | No | Negative |

HCW, healthcare worker; PCR, polymerase chain reaction; Age range was used for participant's privacy and confidentiality.

## Discussion

COVID-19 seroprevalence in asymptomatic staff and patients in Thai community hospitals was higher than hospitals in China (4.5% vs. 2.5%) [3]. Seroprevalence in asymptomatic hospital staff in Thailand was also higher than hospitals in China (3.5% vs. 1.8%) [3] but less than a tertiary hospital in Belgium (3.5% vs. 6.4%) [4]. Asymptomatic patients in Thailand seemed to have higher seroprevalence than China (8.9% vs. 3.5%) [3]. Unlike China and Belgium where the seroprevalences were mostly from positive IgG, our study revealed mostly positive IgM. Comparison with Belgium hospital should be interpreted with caution due to the unknown PCR status of Belgium subjects.

Serological testing provides some crucial epidemiological information and would have been more effective when combined with other diagnostic tests such as PCR. While all participants with positive results from the free-of-charge rapid IgM/IgG test provided in this study were encouraged to get PCR testing, a majority of community hospitals still did not have access to the PCR testing because of both financial and non-financial reasons. Recommendation for PCR testing after positive rapid IgM/IgG test had not fully complied so we did not have information about the participants with positive IgM. With immunoglobulin status and PCR results, we can shape the situation more accurately for both individual and regional views. Hopefully, with this and other vigorous and dedicated studies on antibody status around the globe, serology testing would provide useful information for pandemic control.

## Conclusions

COVID-19 antibody test could detect a substantial number of potential silent spreaders in Thai community hospitals. Antibody testing should be encouraged for mass screening, especially in asymptomatic individuals.

## Data Availability

Data is available from the corresponding author upon request.

## Acknowledgements

We thank Baiya Phytopharm, Thailand for supporting the Baiya Rapid COVID-19 IgG/IgM test kit. The company did not involve in the data analysis, interpretation, and the manuscript preparation.

## Author Contributions

**Conceptualization:** TN, KP, NH

**Data curation:** TN, KC, KP

**Formal analysis:** TN, KP

**Investigation:** TN, KC, KP

**Methodology:** TN, KP, NH

**Project administration:** TN, KP, NH

**Resources:** KP, NH

**Software:** TN, KP

**Supervision:** KP, NH

**Validation:** TN, KC, KP

**Visualization:** TN

**Writing – original draft:** TN, KP

**Writing – review & editing:** TN, KP, KC, NH

